# Glucocorticoid-regulated bidirectional enhancer RNA transcription pinpoints functional genetic variants linked to asthma

**DOI:** 10.1101/2022.11.10.22281906

**Authors:** Sarah K. Sasse, Amber Dahlin, Lynn Sanford, Margaret A. Gruca, Arnav Gupta, Fabienne Gally, Ann Chen Wu, Carlos Iribarren, Robin D. Dowell, Scott T. Weiss, Anthony N. Gerber

**Author notes:** Corresponding Author: Dr. Anthony N. Gerber, Department of Medicine, National Jewish Health, Room K729, 1400 Jackson St, Denver, CO, 80206, USA. Equal contribution (Sarah K. Sasse, Amber Dahlin). **Funding sources**: NHLBI R01 HL109557 (to A.N.G.); NHLBI R01 HL152244 (to A.D.); NIH R01 GM125871 (to R.D.D.). **Conflict of interest disclosure statement**: A.N.G. and S.K.S. own shares in Psammiad Therapeutics; R.D.D. is a cofounder of Arpeggio Biosciences; S.T.W. receives royalties from UpToDate and is on the Scientific Board of Histolix.

## Abstract

Genome-wide association studies of asthma have not explained environmental risk or variable clinical efficacy of glucocorticoids. Bidirectional enhancer RNA (eRNA) transcription is a widespread response to environmental signals and glucocorticoids. Therefore, we investigated whether single nucleotide polymorphisms (SNPs) within dynamically regulated eRNA-transcribing regions contribute to genetic variation in asthma. Through applying multivariate regression modeling with permutation-based significance thresholding to a large clinical cohort, we identified novel associations between asthma and 35 SNPs located in eRNA-transcribing regions implicated in regulating diverse cellular processes relevant to asthma. Functional validation established that *rs258760* (mean allele frequency = 0.34, asthma odds ratio = 0.95; P = 5.04E-03) eliminates an active aryl hydrocarbon receptor (AHR) response element linked to transcriptional regulation of the glucocorticoid receptor gene by AHR ligands commonly found in air pollution. Our findings establish eRNA signatures as a tool for discovery of functional genetic variants and define a novel link between air pollution, glucocorticoid signaling and asthma.

## Introduction

Asthma is a highly prevalent disease for which pathogenesis and treatment responsiveness are governed by complex stochastic interactions between genetic background and the environment^1^. Genome-wide association studies (GWAS) have defined numerous single nucleotide polymorphisms (SNPs) and associated pathways linked to risk of developing asthma^2–4^. For example, a large-scale GWAS of asthma in multiple ethnic populations identified 16 high-confidence SNP associations, annotated to *HLA-DQA1* and *IL1RL1*/*IL18R1*, which have established functional roles for asthma^5^. However, the sum of heritability explained by these SNPs is estimated to account for only 2.5% of the genetic risk of asthma^6–8^. Moreover, the majority of asthma-associated SNPs defined through GWAS are in non-protein coding portions of the genome^9^, and direct regulatory effects of most such SNPs have not been established^10^. These relative weaknesses of GWAS-based approaches are likely due to several underlying factors, including unaccounted effects of environment on asthma risk, linkage disequilibrium, and the very stringent p-values needed to identify significant genetic associations on a genome-wide basis^11^. These drawbacks have also limited the discovery of genetic variants relevant to specific asthma phenotypes, such as responsiveness to therapeutics. For example, clinical heterogeneity in responses to glucocorticoids in asthma is well described, however, relatively few genetic variants have been discovered that directly modify transcriptional responses to glucocorticoids or that directly link glucocorticoid targets to asthma risk^12^.

Specialized regions within the genome that control gene expression, frequently referred to as cis-regulatory elements or enhancers, are enriched for functional genetic variants that are linked to disease heritability^13,14^. As a method to reduce the p-value burden of traditional GWAS and increase the likelihood of identifying putative functional variants, strategies that incorporate enhancer features into genetic association studies have been developed and applied to various diseases^15^. However, enhancers, as defined using standard assays for chromatin accessibility or histone modifications, can span hundreds of kilobases^16^, which impacts resolution and statistical power. Moreover, these approaches often fail to define relationships between enhancers and regulation of specific genes^17^, especially in the context of disease-associated signaling pathways that reshape both enhancer activity and gene transcription.

Sequencing of non-polyadenylated or nascent transcripts has revealed that RNA is transcribed from active enhancers^18,19^, and that concomitant with changes in gene expression, significant changes in enhancer RNA (eRNA) transcription^20,21^ occur in response to a range of stimuli, including steroid hormones, inflammatory signals and environmental pollutants. Although the biologic functions of enhancer RNAs remain poorly understood, eRNA transcription, which can be identified based on sites of bidirectional transcription exclusive of annotated gene transcription start sites, has emerged as a very sensitive marker of enhancer activity^22^. Moreover, discrete sites of RNA polymerase II (RNAPII) loading and transcription initiation within enhancers, which frequently serve as a nidus for the dynamic binding of regulatory transcription factors^19,23^, including the glucocorticoid receptor, can be bioinformatically inferred based on bidirectional transcription signatures^23^. We hypothesized that regions localized to sites of RNAPII loading within eRNA-transcript signatures that change dynamically in response to anti-inflammatory glucocorticoids or pro-inflammatory signals in airway epithelial cells harbor functional genetic variants associated with asthma risk. Here, we combine multivariate logistic regression modeling with permutation-based significance testing to filter SNPs within a large asthma cohort based on localization within dynamic eRNA-transcript signatures, and we perform fine mapping, annotation and functional validation of a subset of these novel asthma-associated SNPs.

## Results

### Localization of SNPs within dynamically regulated enhancers and asthma associations in a large clinical cohort

We previously employed Global Run-on Sequencing (GRO-seq) to analyze nascent RNA transcription on a genome-wide basis in BEAS-2B cells in response to 30-minute treatment with the prototypical anti-inflammatory glucocorticoid, dexamethasone (dex), and/or TNF, a canonical inducer of NFkB signaling, widely used to model therapeutic glucocorticoid cross-talk with inflammatory signals^20^. Revised analysis of these data with updated algorithms (see Methods) defined, exclusive of transcription start sites (TSSs), a set of 1714 unique bidirectional enhancer RNAs (eRNAs) that responded significantly to dex (672 upregulated, 207 downregulated) or TNF (752 upregulated, 83 downregulated), and 137 eRNAs that responded to more than one condition (Supp File 1; p_adj_ < 0.05 vs vehicle). We used Tfit modeling to systematically infer and annotate the sites of RNAPII loading and bidirectional transcription initiation^24^, frequently referred to as Mu (μ), within this eRNA set. To investigate potential relationships between these eRNA-transcribing genomic regions and asthma risk in the non-Hispanic white population within the Genetic Epidemiology Research in Adult Health and Aging (GERA) clinical cohort^5,25^, we leveraged regression modeling, permutation testing and fine mapping approaches^26^ to create an analysis pipeline (Fig 1A) for discovery of genetic variants associated with asthma based on proximity to μ.

**Figure 1.**
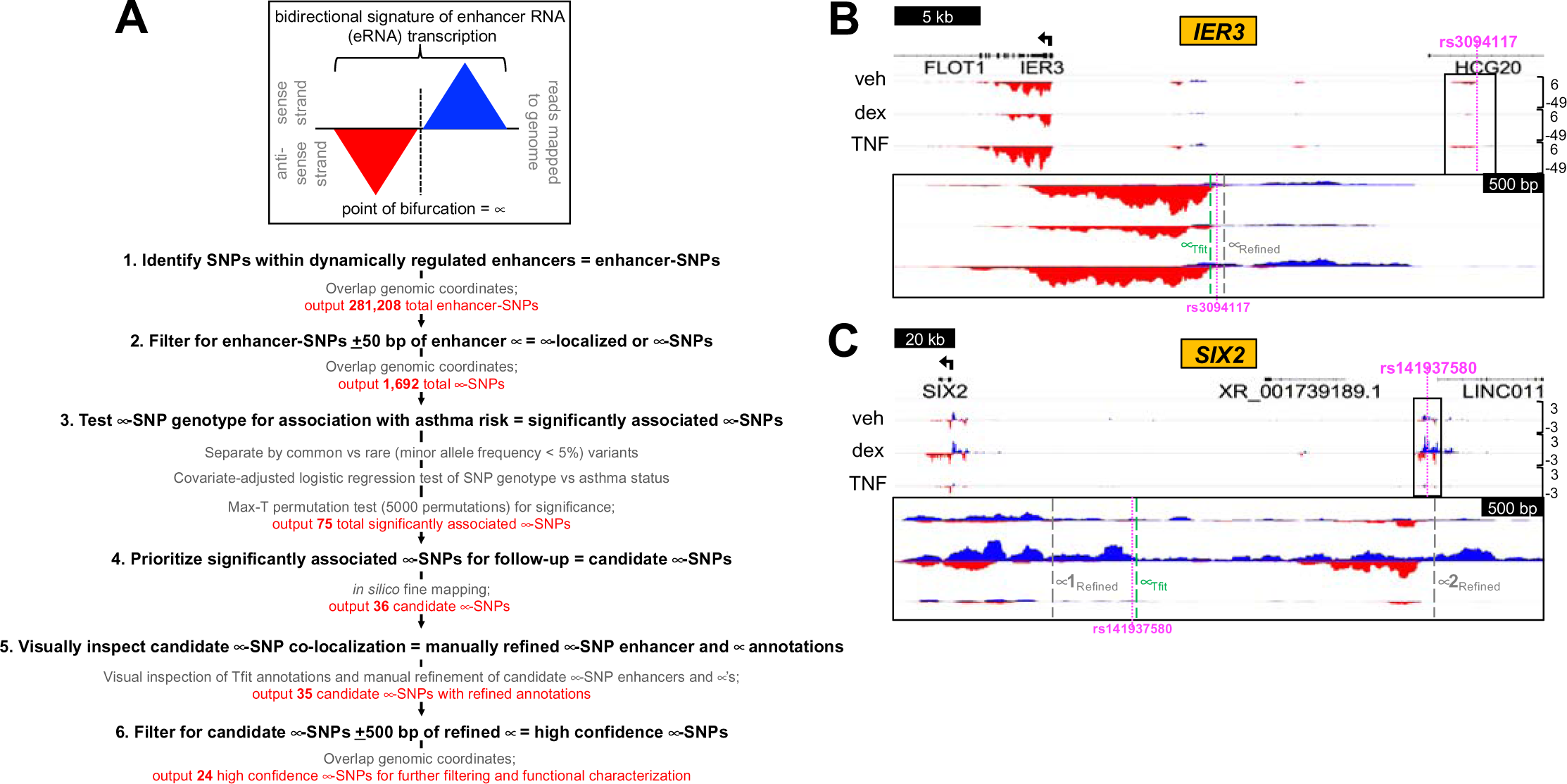
Pipeline for discovery of novel asthma-SNP associations based on proximity to μ and examples of μ-SNP colocalization. **(A)** Schematic of pipeline and filtering approaches to identify high confidence μ-SNPs associated with asthma. (**B-C)** GRO-seq tracks from BEAS-2B cells treated as indicated for 30 min and visualized in the Integrative Genomics Viewer (IGV) genome browser based on counts per million mapped reads (vertical scales). Blue indicates reads annotated to the sense strand while red indicates reads annotated to the antisense strand. The transcription start site (TSS) and direction of transcription are marked by arrows at the top of each screenshot. Magnified regions show the locations of μ originally calculated by Tfit (green) and following manual refinement (gray) relative to the indicated SNP within dynamically regulated enhancers containing high **(B)** or low **(C)** confidence μ-SNPs.

We first screened the GERA dataset to filter SNPs that were annotated within the dynamically regulated enhancer regions, for each treatment separately (dex or TNF). A total of 143,869 SNPs were mapped to dex-responsive enhancer regions and 137,339 were mapped to TNF-responsive enhancer regions. Next, since RNAPII loading sites within eRNA transcribing regions are known to be enriched for functional transcription factor binding motifs^23^, we filtered these enhancer-SNP sets to include only those that were annotated to regions within 50 bp (+/−) of the Tfit-predicted RNAPII loading site, μ (μ-localized SNPs, or μ-SNPs). Within dex-responsive regions, we identified 866 total annotated μ-SNPs (661 upregulated; 205 downregulated), and within TNF-responsive regions we found 826 μ-SNPs (743 upregulated; 83 downregulated). The lower number of μ-SNPs observed within downregulated regions follows the predicted biological function of enhancers, i.e. dex or TNF perturbation are increasingly recognized as directly affecting gene transcription through upregulating enhancer site function^27,28^.

The subsets of μ-SNPs grouped by changes in eRNA transcription with treatment were then tested for association with asthma status using multivariate regression modeling, and the significance of associations was determined using a permutation-based approach implemented in the statistical genetic analysis software, PLINK (v. 1.9)^29^. Across the four treatment groups, representing 1692 total annotated μ-SNPs, 75 μ-SNPs were initially identified as significantly associated with asthma status (25, dex upregulated; 15, dex downregulated; 32, TNF upregulated; 3, TNF downregulated). SNPs within these regions were further characterized using *in silico* fine mapping to investigate possible functional impacts and prioritize candidate hits for follow-up study. Ultimately, this analysis pipeline output 39 significantly associated μ-SNPs with compelling evidence for functional roles based on robustness of association with asthma (Table 1) and proximity to _μ_ (Supp Table 1). Of these, there were 36 unique μ-SNPs, with 3 μ-SNPs identified within an eRNA region that was regulated by both TNF and dex (*rs78553489*, *rs186724791*, and *rs150808839*).

**Table 1.** Initial set of unique μ-SNPs (n=36) significantly associated with asthma risk in GERA cohort, sorted by Permutation P-value.

| SNP | Minor Allele Frequency (GERA) | Odds Ratio (Asthma risk) | SE | L95 | U95 | Differentially Regulated Enhancer Dataset Assoc. with μ-SNP | Permutation Analysis Pval |
| --- | --- | --- | --- | --- | --- | --- | --- |
| rs117861858 | 4.92E-05 | 10.42 | 0.9547 | 1.604 | 67.68 | dex-repressed | 1.13E-04 |
| rs141870749 | 2.95E-05 | 9.824 | 1.473 | 0.5479 | 176.1 | dex-induced | 1.58E-04 |
| rs150872583 | 2.95E-05 | 6.952 | 1.227 | 0.6272 | 77.06 | dex-induced | 1.59E-04 |
| rs3129975 | 2.14E-01 | 1.08 | 0.02 | 1.04 | 1.12 | dex-repressed | 1.65E-04 |
| rs113433891 | 1.97E-05 | 5.073 | 1.421 | 0.3132 | 82.16 | dex-induced | 2.34E-04 |
| rs80028982 | 9.83E-06 | 3.088 | 1.423 | 0.1898 | 50.24 | dex-induced | 2.35E-04 |
| rs149411423 | 2.95E-05 | 13.2 | 1.692 | 0.4791 | 363.5 | dex-induced | 2.36E-04 |
| rs187844646 | 1.97E-05 | 12.58 | 1.554 | 0.5983 | 264.5 | dex-induced | 2.36E-04 |
| rs141937580 | 1.97E-05 | 5.296 | 1.974 | 0.1107 | 253.4 | dex-induced | 2.40E-04 |
| rs182978446 | 1.97E-05 | 10.79 | 1.515 | 0.5546 | 210 | dex-induced | 2.41E-04 |
| rs186639917 | 1.57E-04 | 5.43 | 0.6348 | 1.565 | 18.84 | dex-induced | 1.61E-03 |
| rs78553489 | 2.07E-04 | 4.121 | 0.5667 | 1.357 | 12.51 | TNF-repressed | 4.71E-03 |
|  |  |  |  |  |  | dex-induced | 6.21E-03 |
| rs258760 | 3.40E-01 | 0.95 | 0.02 | 0.92 | 0.99 | dex-repressed | 5.04E-03 |
| rs189641630 | 6.59E-04 | 2.155 | 0.2877 | 1.226 | 3.788 | dex-induced | 6.00E-03 |
| rs141243502 | 2.56E-04 | 0.1491 | 0.7984 | 0.03118 | 0.7132 | dex-induced | 6.50E-03 |
| rs12506479 | 2.71E-01 | 1.05 | 0.02 | 1.01 | 1.09 | TNF-induced | 6.58E-03 |
| rs114363579 | 3.34E-04 | 2.464 | 0.3741 | 1.184 | 5.129 | dex-repressed | 9.30E-03 |
| rs61289882 | 2.08E-01 | 1.05 | 0.02 | 1.01 | 1.09 | TNF-induced | 9.38E-03 |
| rs73731205 | 1.42E-03 | 0.5454 | 0.2431 | 0.3387 | 0.8783 | dex-repressed | 1.02E-02 |
| rs76528672 | 7.87E-05 | 5.51 | 0.7271 | 1.325 | 22.92 | dex-repressed | 1.06E-02 |
| rs76717004 | 3.09E-03 | 0.6658 | 0.161 | 0.4856 | 0.9128 | dex-repressed | 1.15E-02 |
| rs58128597 | 7.54E-02 | 0.93 | 0.03 | 0.87 | 0.98 | TNF-repressed | 1.33E-02 |
| rs78292063 | 1.60E-03 | 1.551 | 0.1835 | 1.082 | 2.222 | dex-induced | 1.41E-02 |
| rs57722273 | 2.38E-03 | 1.431 | 0.1563 | 1.053 | 1.943 | dex-induced | 2.13E-02 |
| rs186724791 | 1.57E-04 | 0.07542 | 1.231 | 0.006753 | 0.8423 | dex-induced | 2.24E-02 |
|  |  |  |  |  |  | TNF-induced | 2.24E-02 |
| rs1454917 | 3.40E-01 | 1.04 | 0.02 | 1 | 1.07 | dex-repressed | 2.83E-02 |
| rs186542091 | 8.92E-03 | 0.8214 | 0.08997 | 0.6887 | 0.9799 | dex-repressed | 3.29E-02 |
| rs1409773 | 6.30E-02 | 1.07 | 0.03 | 1 | 1.14 | TNF-induced | 3.64E-02 |
| rs3094117 | 2.72E-01 | 1.04 | 0.02 | 1 | 1.07 | dex-repressed | 3.69E-02 |
| rs3924229 | 8.19E-02 | 0.94 | 0.03 | 0.89 | 1 | dex-induced | 3.92E-02 |
| rs78124271 | 1.48E-03 | 1.504 | 0.2011 | 1.014 | 2.23 | dex-induced | 4.03E-02 |
| rs188187807 | 1.77E-04 | 2.729 | 0.5261 | 0.9733 | 7.652 | dex-induced | 4.20E-02 |
| rs150808839 | 1.17E-02 | 0.856 | 0.07654 | 0.7368 | 0.9945 | dex-induced | 4.29E-02 |
|  |  |  |  |  |  | TNF-induced | 4.48E-02 |
| rs60249091 | 1.82E-01 | 1.04 | 0.02 | 1 | 1.08 | TNF-induced | 4.33E-02 |
| rs6018492 | 8.64E-02 | 0.95 | 0.03 | 0.89 | 1 | dex-induced | 4.78E-02 |
| rs184589179 | 2.72E-03 | 0.7231 | 0.1632 | 0.5251 | 0.9956 | dex-induced | 4.98E-02 |

As an initial step in validating the relevance of this μ-SNP set in relationship to asthma, searches of the NHGRI-GWAS catalog, ClinVar, and PubMed databases were performed. None of the SNPs were reported as previously associated with asthma by GWAS, suggesting these could represent novel asthma associations based on our methods. Since our analysis is not driven by identifying GWAS signals, but rather as a tool to define functional variants and associated target genes, we alternately interrogated for prior associations at the gene level, specifying overlapping or nearest genes to each μ-SNP as the target. Using published data available in the NHGRI-GWAS catalog and a literature search of PubMed, we identified multiple associations with asthma and/or asthma treatment response, including *SLC16A12* (*rs78124271*), *CXCL8* (annotated as the closest gene downstream of *rs12506479*), *DUSP4* (annotated as the closest gene downstream of *rs186639917* and *rs80028982*), and *CLDN1* (annotated as the closest gene downstream of *rs60249091*). In aggregate, based on proximity, this preliminary analysis revealed that at least 47% (17/36) of our reported loci were previously related to respiratory disease and relevant immune response phenotypes. These data are summarized in Supp Table 2.

### Refined mapping and analysis of _μ_-SNP associations

Confidence in predicting RNAPII loading sites based on Tfit-annotated enhancers depends on the complexity and magnitude of the bidirectional eRNA signature. To focus on SNPs that co-localize with high confidence sites of RNAPII loading (μ), and are thus more likely to be directly associated with enhancer function, we visually inspected the GRO-seq tracks within each enhancer region of interest to manually annotate and refine the bidirectional calls (Supp File 2). One non-unique _μ_-SNP (*rs150808839*) was excluded here due to lack of visual confirmation of a clear bidirectional signature in its surrounding region. The remaining 35 regions were passed to a custom script that calculates the predicted location of μ based on each manually refined bidirectional annotation. A total of 24 _μ_-SNPs (69%) were located within 500 bp (+/−) of the refined _μ_ sites; these were classified as high confidence _μ_-SNPs and prioritized for further analysis (Supp Table 3). Examples of high confidence and lower confidence sites of _μ_-SNP co-localization are depicted in Figs 1B-C, respectively. Notably, of the 24 high confidence _μ_-SNPs, 22 (92%) resided in enhancers that, based on similar transcriptional patterns in response to dex and/or TNF (Supp Files 3-4; p_adj_ < 0.05 vs vehicle), could be linked to regulation of at least one proximal gene within a one megabase (Mb) radius (Supp Table 3). Gene functions linked to individual SNPs in this manner encompass signaling processes (e.g. *BMP1*^30^ and *PLK2*^31^), ubiquitin ligase activity (*HERC4*^32^) and transcriptional regulation (*CEBPB*^33^).

To further assess the regulatory function of the high confidence _μ_-SNP regions, we cloned ∼450-1100 bp spanning 9 of the high confidence _μ_-SNP regions into a minimal promoter luciferase reporter (Supp Table 4). We assayed responses of these reporters to dex and TNF in BEAS-2B cells in comparison to two control glucocorticoid-responsive reporters (p*FKBP5*, p*TNFAIP3*) and two reporters derived from low confidence _μ_-SNP regions (p*COL8A1*, p*CFLAR*). We found that 7 of the 9 high confidence enhancer reporters (78%) exhibited dynamic changes in activity (Fig 2A) consistent with regulation of the corresponding endogenous eRNAs (Fig 2B; representative examples). These data indicate that the high confidence _μ_-SNPs are likely to confer biologically relevant regulatory function.

**Figure 2.**
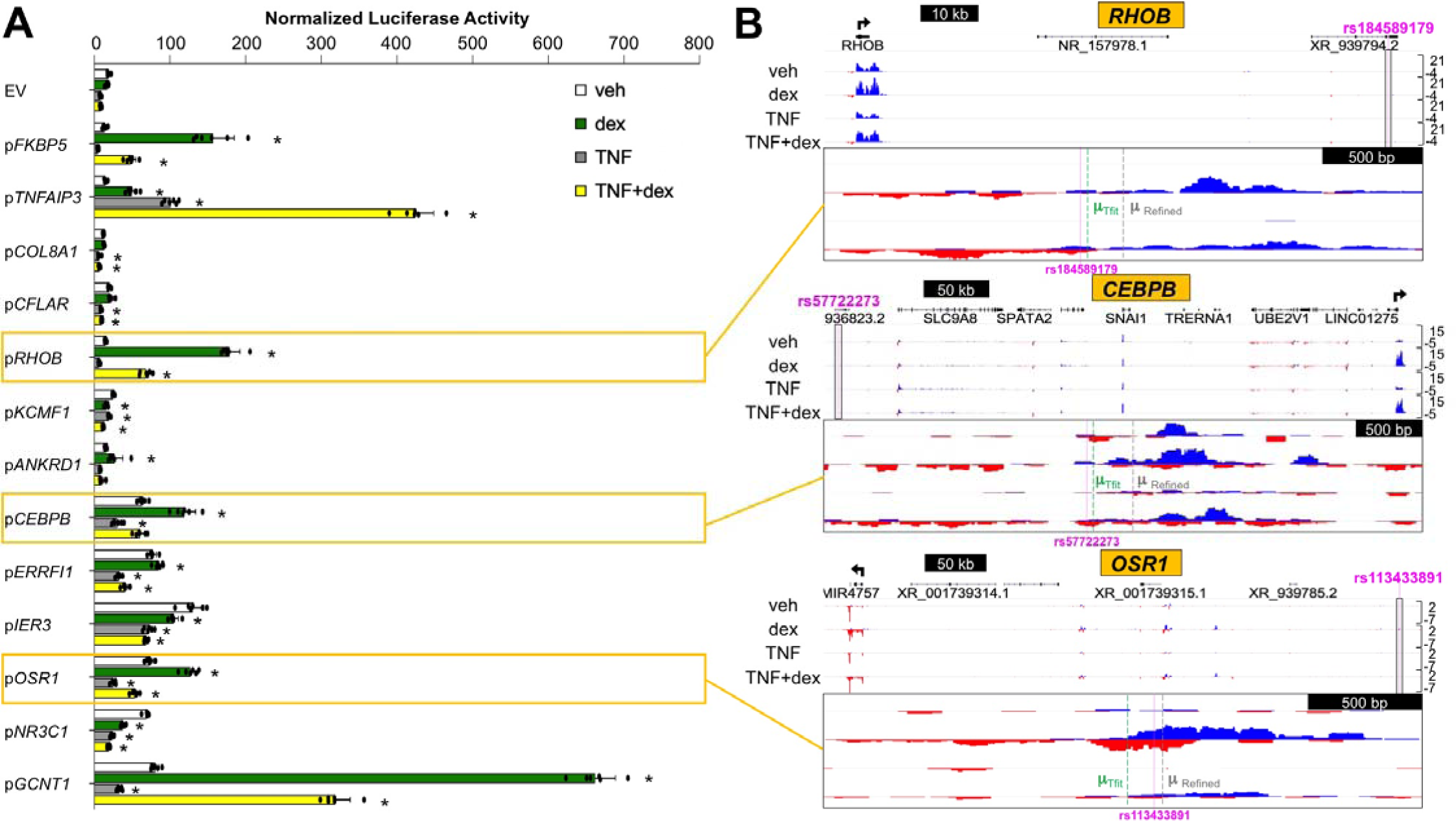
High confidence μ-SNP regions recapitulate dynamic eRNA regulation patterns in reporter assays. **(A)** Mean (±SD) normalized luciferase activity of indicated enhancer reporter constructs or empty vector (EV) control in BEAS-2B cells treated as indicated for 8 hr. p*FKBP5* and p*TNFAIP3* were included as canonical positive controls for dex and TNF responses. *p<0.05 vs same reporter+veh; one-way ANOVA corrected for multiple comparisons **(B)** GRO-seq tracks, as described for Figure 1, for select interrogated by enhancer reporter assay.

### Functional analysis characterizes novel SNPs relevant to GR signaling

To identify high confidence _μ_-SNPs that are most likely to exert a functional role in controlling gene expression, we used MatInspector^34^ to annotate canonical transcription factor binding motifs within 30 bp (+/−) of each SNP site. Through comparing motifs identified for both the major and variant alleles, *rs149411423* and *rs258760* emerged as strong candidates for functional regulation. *rs149411423* is a C>T transversion (minor allele frequency (MAF) in GERA = 2.95E-05; Table 1) located in an enhancer region within *PRUNE2* that is ∼175 kb from the TSS of *GCNT1*, which encodes a glycosyltransferase implicated in pulmonary immune responses and allergy^35,36^. As shown in Fig 3A, both *GCNT1* (log_2_FC = 2.19, p_adj_ = 3.93E-09) and the enhancer harboring *rs149411423* (log_2_FC = 2.57, p_adj_ = 0.003) show increased transcription after dex treatment in BEAS-2B cells, whereas *PRUNE2* expression was unchanged (log_2_FC =0.27, p_adj_ = 0.999). GR ChIP-seq analyses in BEAS-2B^20,37^ (Fig 3B, *top*) and primary human airway smooth muscle (HASM) cells^38^ (Fig 3B, *bottom*), another critical effector cell type in asthma, revealed dex-inducible GR occupancy at the *rs149411423*-μ-SNP region. In fact, the *rs149411423*-μ-SNP was by far the most prominent site of inducible GR occupancy across the *GCNT1* locus and spanning the surrounding genomic region, further supporting a regulatory role for this enhancer in the induction of *GCNT1* by glucocorticoids. To further assess putative regulation of *GCNT1* transcription by the *rs149411423*-μ-SNP enhancer region, we generated three-dimensional chromatin contact maps in BEAS-2B cells using Micro-C, which we visualized as arcs connecting interacting genomic regions across the locus of interest (Fig 3C). These data identified three-dimensional contacts between the *rs149411423*-μ-SNP region and the *GCNT1* TSS. Moreover, publicly available Micro-C data generated in human embryonic stem cells^39^ and visualized in the UCSC Genome Browser defined a topologically associated domain (TAD) that contains both the *rs149411423*-μ-SNP and the *GCNT1* TSS (Fig 3D). Taken together, the three-dimensional architecture of the region is consistent with the *rs149411423*-μ-SNP locus regulating expression of *GCNT1*.

**Figure 3.**
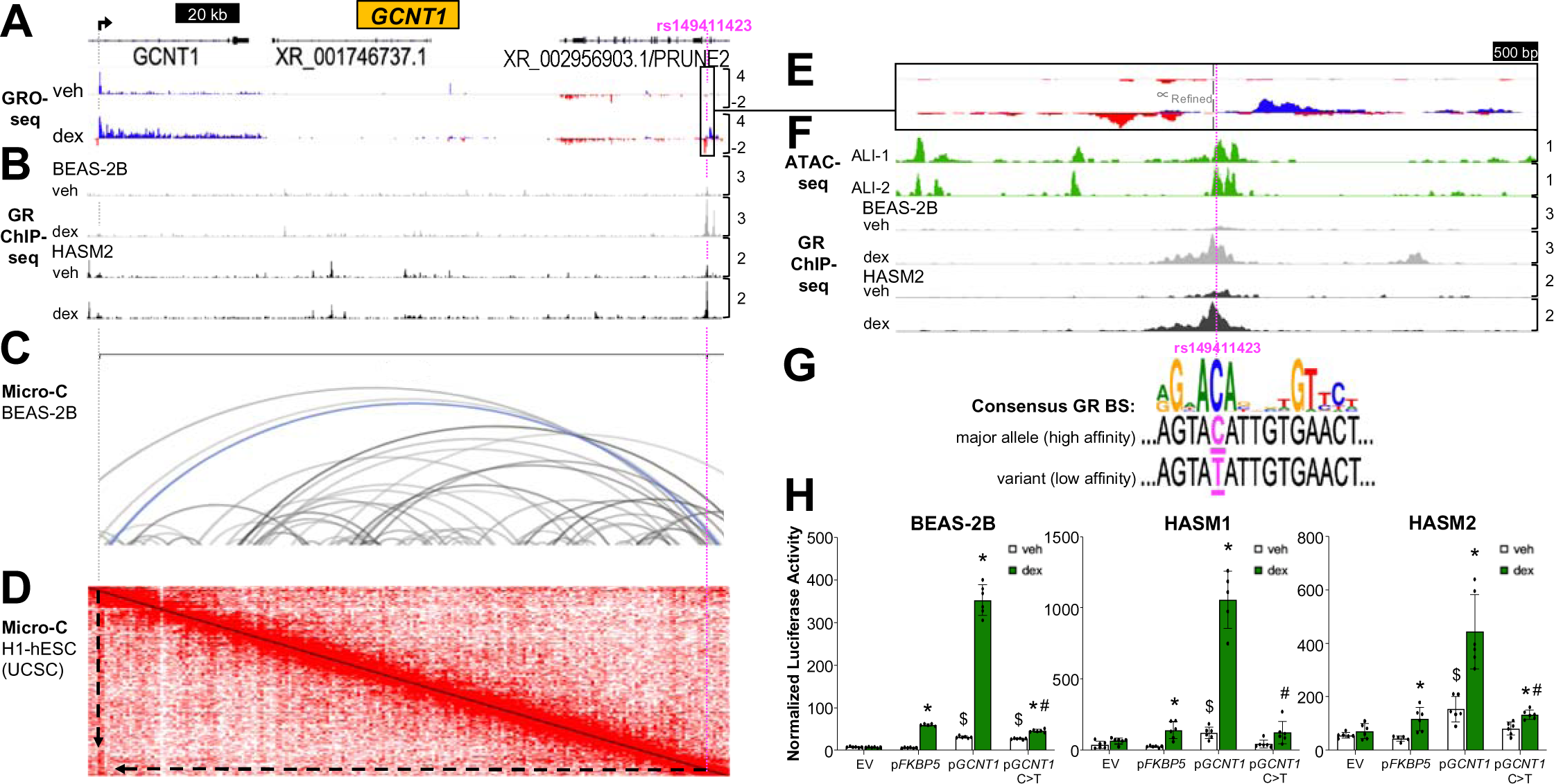
*rs14941123* disrupts a functional glucocorticoid response element that regulates induction of *GCNT1*. **(A)** GRO-seq tracks, as described for Figure 1, for the *GCNT1* locus. Scaling applies through panel (D). **(B)** Glucocorticoid receptor (GR) ChIP-seq tracks from BEAS-2B (gray; *top*) and primary human airway smooth muscle (HASM) cells (black; *bottom*) ±1 hr dex visualized in the IGV browser based on counts per million mapped reads (vertical scales). **(C)** Micro-C three-dimensional chromatin contact map in BEAS-2B cells with arcs connecting 1 kb interacting regions across genomic space; darker color indicates higher frequency of contact; blue indicates contact between regions of interest (*rs14941123* and *GCNT1* TSS). Arc height is proportional to distance between contacts. **(D)** Heat map indicating frequency of three-dimensional physical contacts in publicly available Micro-C data generated in human embryonic stem cells^39^ and visualized in the UCSC Genome Browser; arrows indicate high frequency of contact between regions of interest (*rs14941123* and *GCNT1* TSS). **(E)** Magnified view of GRO-seq tracks within *rs149411423*-μ-SNP region shown in panel (A). Scaling applies through Panel (F). **(F)** Aligned ATAC-seq tracks from primary human airway epithelial cells cultured at air-liquid interface (ALI; green), based on counts per million mapped reads (vertical scales), and magnified view of aligned GR ChIP-seq peaks described in panel (B). **(G)** Consensus binding sequence logo (MatBase) for GR and sequence of match identified by MatInspector within the *GCNT1* enhancer harboring *rs14941123*. **(H)** Mean (±SD) normalized luciferase activity of parent or point mutation *GCNT1* reporter constructs in cells treated ±dex for 8 hr (BEAS-2B) or 24 hr (primary HASM). *p<0.05 vs same reporter+veh; unpaired t-tests corrected for multiple comparisons, ^$^p<0.05 vs EV+veh; one-way ANOVA corrected for multiple comparisons, ^#^p<0.05 vs parent construct+dex; one-way ANOVA corrected for multiple comparisons.

A closer look at the *rs149411423*-μ-SNP enhancer (Fig 3E) illustrates clear overlap of the eRNA bidirectional center with a chromatin accessibility peak in differentiated primary human airway epithelial cells within ATAC-seq data we generated previously^40^ (Fig 3F, *top*), supporting physiological relevance of the region, and the dex-inducible GR ChIP-seq peak observed in both BEAS-2B and HASM cells (Fig 3F, *bottom*). MatInspector analysis of the sequence immediately surrounding *rs149411423* indicated that the transversion site actually resides within a putative glucocorticoid response element (Fig 3G). Through applying a customized position weight matrix we created to predict relative GR binding affinities^41^, we found that the predicted affinity of the sequence containing the major C allele for binding GR is ∼2-fold greater than that containing the variant T allele, suggesting the SNP may affect enhancer function. To assess whether *rs149411423* directly alters GR-regulated transcription of the enhancer, we generated a reporter harboring the variant allele using site-directed mutagenesis and assayed its activity compared to the parent construct with the major C allele in BEAS-2B and primary HASM cells. Relative to the parent construct, the mutant reporter exhibited dramatically attenuated induction by dex in all tested cell types (Fig 3H). In aggregate, our data implicate *rs149411423* as reducing GR-mediated induction of an enhancer that regulates transcription of *GCNT1,* strongly supporting a functional role for this SNP.

The second strong candidate SNP, *rs258760,* is a C>T transversion (MAF in GERA = 3.40E-01; Table 1) within an *ARHGAP26* intron. This genomic location is approximately 200 kb downstream of the TSS for *NR3C1*, which encodes the glucocorticoid receptor. To assess whether the enhancer region harboring *rs258760* is associated with transcriptional control of *ARHGAP26* or *NR3C1*, we compared eRNA transcription patterns to nascent transcription of both genes after dex or TNF+dex treatment. Fig 4A (GRO-seq tracks) illustrates that eRNA transcription from the *rs258760* region is reduced by dex (log_2_FC = −1.8, p_adj_ = 0.035) and TNF+dex treatment (log_2_FC = −2.01, p_adj_ = 0.011). Whereas *ARHGAP26* transcription is increased by dex (log_2_FC = 0.76, p_adj_ = 0.035) and TNF+dex (log_2_FC = 1.41, p_adj_ = 1.04E-07), *NR3C1* transcription is significantly reduced by treatment with TNF+dex (log_2_FC = −1.36, p_adj_ = 0.0008). As no other gene within a 1 Mb window of the *rs258760* region showed significant expression changes with either dex or dex+TNF treatment, these data suggest that *rs258760* resides in a regulatory element that controls expression of *NR3C1*. Further supporting this notion, in our published analysis of nascent transcriptional responses to wood smoke particles (WSP) using Precision Run-on Sequencing (PRO-seq), eRNA transcription from the same +200 kb enhancer was significantly increased after 30 minutes of WSP exposure in BEAS-2B cells (log_2_FC = 1.99, p_adj_ = 6.47E-62), with a congruent increase in transcription of *NR3C1* (log_2_FC = 0.90, p_adj_ = 9.66E-43) but not *ARHGAP26* (log_2_FC = −0.11, p_adj_ = 0.071; Fig 4A, PRO-seq tracks). Together, these data strongly suggest that the enhancer harboring *rs258760* controls transcription of *NR3C1*, rather than *ARHGAP26*. This μ-SNP region also aligns with an ATAC-seq peak in patient-derived airway epithelial cells^40^, providing additional support for the physiologic relevance of this enhancer (Fig 4B). Furthermore, MatInspector analysis of transcription factor binding motifs in this region in the context of the major C vs variant T alleles revealed that the C allele is a critical nucleotide within a canonical binding motif for the aryl hydrocarbon receptor (AHR). This AHR motif is lost in the context of the T allele (Fig 4C), raising the possibility that AHR ligands, which are found at significant concentrations in wood and wildfire smoke^42–44^, control the activity of this enhancer in an allele-specific manner.

**Figure 4.**
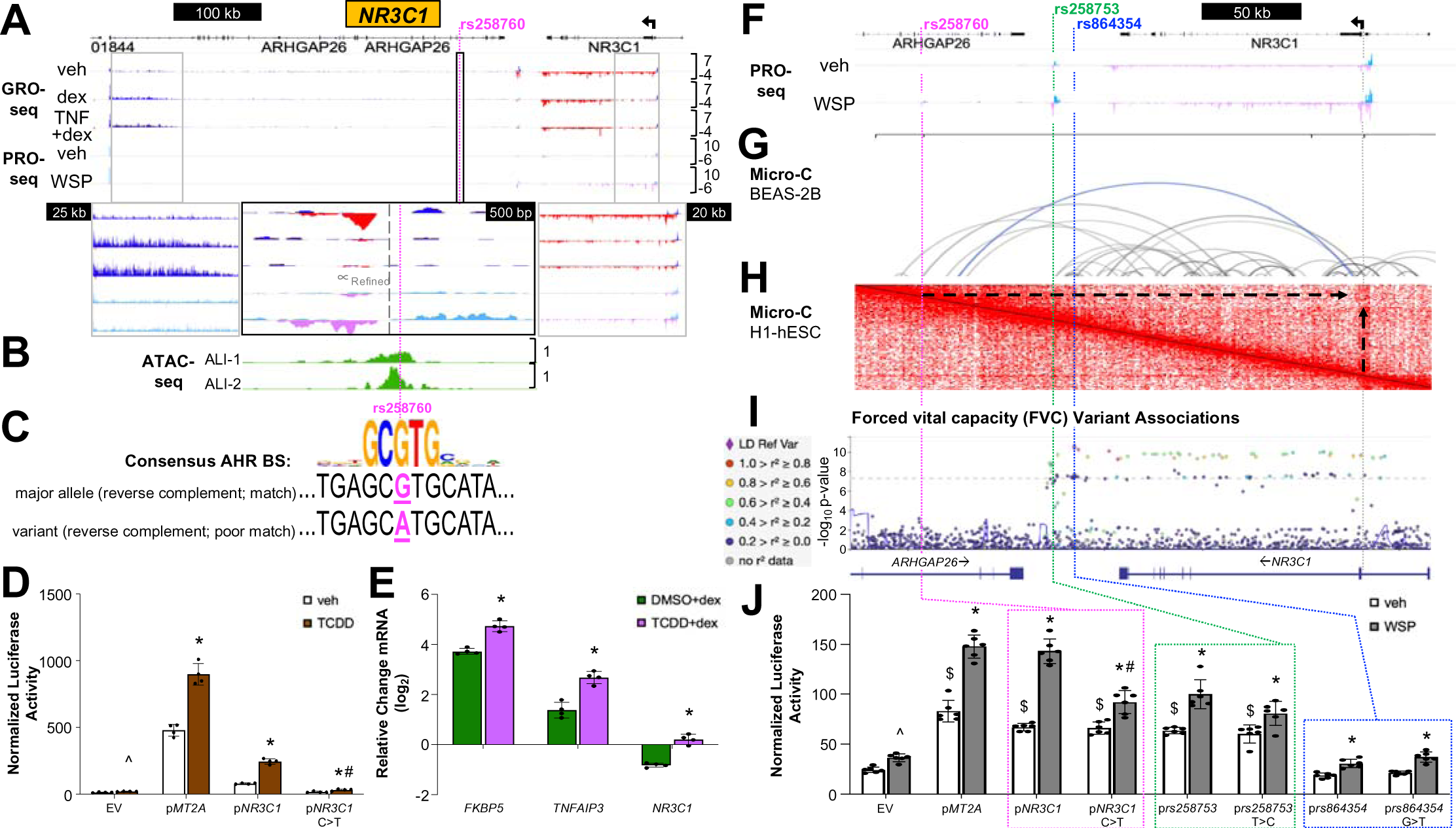
Expression and function of *NR3C1* are regulated by aryl hydrocarbon receptor (AHR) ligands through an AHR response element that is lost in the context of the *rs258760* variant allele. **(A)** IGV screenshots of BEAS-2B GRO-seq tracks aligned with PRO-seq tracks from BEAS-2B cells ±30 min wood smoke particles (WSP) at the *NR3C1* locus. PRO-seq reads annotated to the sense strand are light blue while those annotated to the antisense strand are light purple. Scaling applies through panel **(B)**. **(B)** Aligned ATAC-seq tracks from primary human airway epithelial cells cultured at ALI. **(C)** Consensus binding sequence logo (MatBase) for the aryl hydrocarbon receptor (AHR) and sequence of match identified by MatInspector within the *NR3C1* enhancer containing *rs258760*. **(D)** Mean (±SD) normalized luciferase activity of parent or point mutation *NR3C1* reporter constructs in BEAS-2B cells treated ±TCDD for 8 hr; p*MT2A* served as a positive control for TCDD responsiveness. ^p<0.05 vs EV+veh; AHR ligands consistently induce slight yet statistically significant activity from the empty vector construct; one-way ANOVA corrected for multiple comparisons, *p<0.05 vs same reporter+veh; unpaired t-tests corrected for multiple comparisons, ^#^p<0.05 vs parent construct+TCDD; one-way ANOVA corrected for multiple comparisons. **(E)** qPCR analysis of dex-mediated gene regulation following 4 hr TCDD pre-treatment in BEAS-2B cells. Bars depict mean (±SD) C_T_ values on a log_2_ scale relative to DMSO+veh-treated cells. *p<0.05 vs same gene + DMSO+dex; unpaired t-tests corrected for multiple comparisons. **(F)** Magnified view of BEAS-2B WSP PRO-seq tracks within *ARHGAP26-NR3C1* locus shown in panel (A). Scaling applies through panel (I). **(G)** BEAS-2B Micro-C contact map as described for Fig 3C, with blue indicating contact between regions of interest (*rs258603* and *NR3C1* TSS). **(H)** Human embryonic stem cell Micro-C interaction heat map^39^ as described for Fig 3D, with arrows indicating high frequency of contact between regions of interest (*rs258760* and *NR3C1* TSS). **(I)** Visualization of SNPs by GWAS p-values for associations with lung forced vital capacity (FVC)^47^ across the chr5:143,100,000-143,400,000 (*NR3C1*) topologically-associated domain (TAD) in the Lung Disease Knowledge Portal; the strength of association of SNPs in linkage disequilibrium with the lead FVC GWAS SNP in the region (*rs864354*) is indicated by the color scale on the left. **(J)** Mean (±SD) normalized luciferase activity of parent or point mutation *NR3C1* reporter constructs in BEAS-2B cells treated ±WSP for 8 hr. ^p<0.05 vs EV+veh; one-way ANOVA corrected for multiple comparisons, ^$^p<0.05 vs EV+veh; one-way ANOVA corrected for multiple comparisons, *p<0.05 vs same reporter+veh; unpaired t-tests corrected for multiple comparisons, ^#^p<0.05 vs parent construct+WSP; one-way ANOVA corrected for multiple comparisons.

To determine whether AHR and *rs258760* regulate activity of this presumptive *NR3C1* enhancer, we used site-directed mutagenesis to create a reporter with the minor, asthma-protective T allele, and assayed its activity compared to the parent construct in BEAS-2B cells. As illustrated in Fig 4D, while the parent C allele reporter shows robust induction following exposure to the dioxin, TCDD, a potent AHR ligand responsible for toxic effects of Agent Orange^45^, this induction was markedly reduced in the mutant reporter harboring the minor T allele. Thus, *rs258760* eliminates a functional AHR response element linked to regulating expression of *NR3C1* in airway epithelial cells in response to AHR ligands. Further, pre-treatment of BEAS-2B cells with TCDD prior to dex exposure potentiates induction of canonical GR transcriptional targets (Fig 4E), such as *FKBP5*^46^. These findings implicate the major C allele, which is associated with increased risk of asthma, as integral to an active AHR response element controlling a functional AHR-NR3C1 signaling axis in airway epithelial cells.

To further probe the relevance of the *rs258760*-μ-SNP in controlling *NR3C1* transcriptional regulation, we visualized our BEAS-2B Micro-C data across this region, and found evidence for three-dimensional physical contacts between the *rs258760*-μ-SNP locus and the *NR3C1* TSS (Fig 4F, G). These findings were corroborated by Micro-C data generated in human embryonic stem cells^39^ (Fig 4H), indicating that the *rs258760*-μ-SNP and the *NR3C1* TSS reside within a TAD roughly encompassing chr5:143,100,000-143,400,000 (hg38). To determine whether *rs258760* and/or other SNPs within the chr5:143,100,000-143,400,000 TAD have been independently associated with measures of lung function relevant to asthma, we used the Lung Disease Knowledge Portal (https://lung.hugeamp.org/) to visualize the TAD in genomic space along the x-axis relative to GWAS p-values for associations with lung forced vital capacity (FVC)^47^, which has been previously defined as relevant to air pollution and asthma^48–51^, on the y-axis (Fig 4I). While *rs258760* did not achieve statistical significance, there were significant associations between a number of other SNPs within the chr5:143,100,000-143,400,000 TAD and FVC, suggesting genetic variation within this TAD influences lung function. Of these, the GWAS-defined lead variant in the region, *rs864354* (FVC, GWAS p-value = 3.00E-11), lies within a genomic area that has no features of enhancer activity in our nascent sequencing data (see Fig 4F). In contrast, rs*258753* (FVC, GWAS p-value = 1.74E-9) is located within an intergenic enhancer in which eRNA transcription increases in response to WSP (see Fig 4F). To further explore putative functional relevance of *rs864354* and rs*258753,* we created luciferase reporters driven by the ∼500-800 bp genomic regions spanning each transversion site, and used site-directed mutagenesis to introduce the relevant variant allele into each reporter context. We directly compared the activities of these reporters to the *rs258760*-μ-SNP reporter series in response to WSP treatment (Fig 4J). The reporter spanning the *rs864354* SNP did not exhibit appreciable activity relative to the empty vector control, suggesting the region is not regulatory and the SNP is not likely a functional variant. In contrast, the *rs258753* reporter showed basal enhancer activity that increased in response to WSP, though not as robustly as the *rs258760*-μ-SNP reporter. And while the *rs258753* variant allele reduced responses to WSP relative to the parent construct, the difference was not statistically significant. In aggregate, these data implicate genetic variation in regulatory elements for *NR3C1* that respond to WSP as influencing asthma risk and lung function, and establish a novel connection between asthma, combustion-derived pollutants, and control of *NR3C1* expression.

## Discussion

Nascent transcript sequencing is a powerful method for quantifying enhancer activity and has been applied previously to study eQTLs^52,53^, yet had not been systematically leveraged to identify novel SNP-disease associations. Our results indicate that constraining genetic association analysis based on SNP proximity to bidirectional eRNA transcription initiation sites can facilitate discovery of genetic variants that influence asthma risk, including the identification of disease-associated rare variants, which are difficult to associate with disease using traditional GWAS. SNP filtering and analysis based on nascent eRNA transcription signatures offers other unique advantages including enrichment for functional regions within enhancers, which can span thousands of base pairs when defined using other methods, and facilitating the identification of presumptive target genes for specific SNP-enhancer regions. Moreover, although we focused here on SNPs within eRNA-centered RNAPII loading regions that respond dynamically to glucocorticoids or TNF, this approach is generalizable to eRNAs regulated by other stimuli relevant to asthma, such as Type II cytokines or particulate pollution. Thus, our data provide a systematic method for discovery of functional variants that influence asthma or other airway disease risk in the context of specific endotypes or environmental exposures.

A number of elements of our experimental approach were empirically based on principles of enhancer biology and also designed to address specific limitations of current methodologies. For example, as gene TSSs are well annotated by many methods^54^, and SNPs located at TSSs do not pose an assignment problem in which the identification of the target gene for a putative regulatory SNP is uncertain, we specifically excluded sites of bidirectional transcription that overlap with gene TSSs from our filtering pipeline. In addition, since sites of bidirectional eRNA transcription are enriched for TF binding^19,23^, we used narrow 100 bp regions centered on bidirectional eRNA signatures to filter SNPs prior to permutation analysis. This also served to limit the genomic search space, thereby increasing statistical power. However, when we refined the mapping of bidirectional eRNA transcription in relationship to high priority SNPs identified through our pipeline, we expanded the constraint for μ-SNP proximity to 1 kb regions centered on each refined μ, which better reflects the average size of typical enhancers^55^. Future investigations are needed to determine the optimal size for initial filtering in relationship to μ and to identify centers of bidirectional transcription more efficiently and using scalable bioinformatics methods, with a goal of obviating the manual refinement step we used in this work. Finally, we used congruent transcriptional responses between eRNA-transcribing regions and genes located within a 1 Mb region to empirically assign relevant enhancer-gene interactions, with corroborative support provided by chromatin conformation data. This system expands on methods that have used cap analysis gene expression data from different cell types^56,57^, which can also define enhancer activity, to facilitate enhancer-promoter assignments, and our 1 Mb size window for assignments is derived from these and other data^58^. A key strength of our system in relationship to defining functional SNP-gene-disease relationships is the use of dynamic eRNA and gene transcription data in response to disease-relevant stimuli, rather than reliance on static expression comparisons across different cell types^59^. Genomic editing using CRISPR-based approaches to directly assess functional interactions between a subset of our putative enhancer-reporter pairs would further validate our approach, although enhancer redundancy can complicate such efforts^60^.

Two variants, *rs149411423* and *rs258760,* directly illustrate the effectiveness of our experimental design for discovery of functional SNPs and extend our understanding of GR signaling in the context of asthma risk. *rs149411423,* a high risk, low frequency allele, disrupts a functional GRE that regulates *GCNT1*, a glycosyltransferase linked to IgE levels, immune signaling and the pulmonary immune response to infection^35,61,62^. Thus, although it remains to be determined whether *rs149411423* directly influences the therapeutic effects of GCs in asthma, we elucidate a novel and physiologically plausible GR-GCNT1 axis that is relevant to asthma risk. Interestingly, *rs149411423* was present at very low minor allele frequencies in all populations genotyped in GERA, but was reported at a notably higher frequency in the African-American population genotyped in the 1000 Genomes Project (MAF in 1000 Genomes = 0.0106). Studies that utilize multi-ethnic cohorts with sufficient sample sizes to provide the statistical power to detect robust associations with asthma are needed to further clarify the roles of the variants we identified in this study.

We also discovered a genetic variant, *rs258760*, a low risk, high frequency allele that regulates expression of the glucocorticoid receptor itself in relationship to asthma risk. In this case the variant allele is the most common allele, and minor allele frequencies were again found to vary by ethnicity in other multi-ethnic cohorts such as 1000 Genomes: (*rs258760* MAF in 1000 Genomes = 0.34 for European, 0.117 for African-American and 0.008 for East Asian populations). The more common C allele, associated with increased risk of an asthma diagnosis, is a required nucleotide within a functional binding site for AHR. Our data implicate this AHR response element in controlling increased transcription of airway epithelial GR in response to AHR ligands, which are present at physiologically relevant concentrations in particulate matter air pollution^63,64^. Our results are supported by evidence that *rs258760* and *NR3C1* reside within a TAD and physically associate with each other in three dimensions. Moreover, within this TAD, a second SNP, *rs258753*, which achieved genome-wide significance in a GWAS examining lung forced vital capacity (FVC), is located within a distinct wood smoke-induced enhancer. However, this SNP was not predicted to alter binding site preferences for AHR, and accordingly, our data show that only *rs258760* significantly influenced enhancer responses to wood smoke. Thus, we establish an unexpected genetic link between combustion-related air pollution, such as wood smoke, regulation of GR expression, and lung function. Paradoxically, as the loss of a functional AHR element caused by the *rs258760* variant allele appears to decrease the risk of asthma, our data support a model in which local inflammatory repression via GR in response to pollution may have deleterious effects. This raises the possibility that use of inhaled steroids in the context of air pollution, a problem of increasing importance given the malign effects of climate change on air quality^65–67^, may entail unappreciated risks dependent on genetic context. While further exploration and validation are underway, particularly in appropriately powered multi-ethnic cohorts, in aggregate, we have defined novel genetic links between GR signaling and asthma risk, and we have established a powerful new application of eRNA transcription signatures for discovery of SNP-disease associations.

## METHODS

### Nascent transcription and cistromic datasets

We previously published the BEAS-2B GRO-seq nascent transcript dataset^20^ used in the current study with linked Gene Expression Omnibus (GEO) accession number GSE124916 (30 min datasets). Duplicate samples for four treatment conditions [vehicle (ethanol), TNF (20 ng/ml), dexamethasone (100 nM) or TNF+dex] at two time points (10 or 30 minutes) were sequenced in the original study. Raw sequencing data in fastq format as well as non-normalized, strand-differentiated coverage files of hg38-mapped reads in bedGraph format are available for download under this accession series. For the current study, computational analysis to identify regions with bidirectional transcriptional activity (eRNAs) was performed exactly as described^20^ with the following modification: a newly available version (https://github.com/Dowell-Lab/FStitch) of the Fast Read Stitcher (FStitch) program (v. 1.1.1), with improved efficiency and accuracy impacts on downstream processing, was used for file pre-processing prior to application of Tfit (v. 1.0) to annotate the bidirectional eRNA signatures. Revised eRNA annotations were then subjected to differential expression analysis using DESeq2 to identify differentially transcribed eRNAs (Supp File 1) and their inferred sites of RNAPII loading (μ). The GEO accession number for the primary human airway epithelial cell ATAC-seq data (Norm-1 and Norm-2 datasets), described previously^40^, is GSE157606. BEAS-2B ChIP-seq data were published previously^37^, with GEO accession number GSE79803. HASM ChIP-seq data were also described previously^38^, with GEO accession number GSE95632. Here, to create the screenshots depicted in Figure 3, we remapped the BEAS-2B and HASM ChIP-seq datasets to hg38 using our standardized ChIP-seq pipeline. Briefly, reads are trimmed for adapters, minimum length, and minimum quality using the bbduk tool from the BBMap Suite (v. 38.05). Trimmed reads are then mapped to hg38 using hisat2 (v. 2.1.0). Resulting SAM files are converted to sorted BAM files using samtools (v. 1.9) and to bedGraph coverage format using genomeCoverageBed from the BEDTools suite (v. 2.29.2). Read coverage is normalized to reads per million mapped using a custom python script and files converted to TDF format using igvtools (v. 2.5.3) for visualization in IGV. The BEAS-2B 30-minute wood smoke particle exposure PRO-seq data depicted in Figure 4 were published previously^68^, with GEO accession number GSE167371.

### Genomic data

We utilized genotype (SNP) and phenotype information (asthma diagnosis, age, sex, body mass index (BMI), length of follow up, smoking status (current, former, never), and the first six ancestral principal components) from the Genetic Epidemiology Research on Adult Health and Aging (GERA) cohort, which includes over 110,000 subjects (68,623 total asthma cases across four ethnic groups) with extensive electronic medical records (EMR) information and genome-wide SNP genotypes (>8 million typed and imputed markers). As GWAS-based associations of genetic risk factors with disease susceptibility can vary significantly by ethnic group within large multi-ethnic cohorts such as GERA^5^, we limited the current study to a subset of the cohort with the highest percentage of asthma cases (16,274 non-Hispanic white asthmatic cases and 38,269 controls).

### μ-SNP co-localization and statistical association with asthma risk

Unless otherwise indicated, R statistical computing software and PLINK^29^ (v. 1.9; http://pngu.mgh.harvard.edu/purcell/plink) were used for all analyses.

#### Statistical methods

To identify genetic predictors for asthma that were also present in dynamically regulated enhancer regions, we first screened the GERA subset (54,543 samples) to filter SNPs that overlapped the Tfit-annotated enhancer regions significantly regulated by dex or TNF (Supp File 1), for each treatment separately. Next, we filtered these SNP sets to include only those that were annotated to regions within 50 bp (+/−) of the Tfit-predicted RNAPII loading site (μ), referred to as μ-SNPs. After sorting by minor allele frequency (MAF), μ-SNPs were categorized as common (MAF ≥ 0.05) or rare (MAF < 0.05) variants and tested independently. Tests for association of additive genotype (categorical variable) with asthma status (dichotomized outcome variable) were conducted using multivariate logistic regression models, adjusted for covariates (listed in the “Genomic data” section above). The significance of associations was determined using a permutation-based approach. One method to improve feature prediction is to combine model-based approaches with permutation. Since each set of dynamically regulated enhancer regions contained relatively few (<1000) SNPs genotyped in GERA, yielding a small number of tests, far fewer SNPs were interrogated overall as compared to a GWAS, which tests millions of SNPs. In this instance, multiple correction methods traditionally applied in GWAS (e.g. Bonferroni) are overly conservative, or over-correct for Type I error. In contrast to the Chi-square test, permutation-based approaches are appropriate for analysis of rare alleles or smaller numbers of samples and preserve the correlational structure between SNPs, requiring a less stringent multiple test correction threshold. Max(T) permutation was applied with 5000 permutations to generate two sets of empirical (pointwise and familywise) P values. The max(T) permutation approach is advantageous for candidate SNP-based approaches, as it is typically sufficient to perform a much smaller number of tests. Following permutation and multiple testing correction using a false-discovery rate (FDR) threshold of 0.05, associations were prioritized based on their level of significance. PLINK v. 1.9 was used to conduct permutation-based analyses while R was employed for SNP filtering, analysis of summary statistics, and prioritization.

#### Fine mapping and candidate SNP selection

An *in silico* fine mapping approach was applied using a web-based annotation tool, SNPnexus^69^ (v. 4; https://www.snp-nexus.org/v4/), to determine the SNP genomic localization, predicted functions (coding, non-coding, etc.), predicted consequences on protein/gene expression or function, relationship to regulatory elements (e.g. transcription factor motifs, CpG windows), and impact on structural variation (e.g. CNVs). Prior reports of associations with asthma, lung function measures, glucocorticoid response or related phenotypes were also determined through literature search using the NCBI PubMed database. After fine mapping, SNPs were grouped according to whether their respective enhancer regions were up- or down-regulated and compared across groups. The resulting set of SNPs with the strongest predictive evidence for causality and closest proximity to _μ_ were prioritized for functional characterization *in vitro*.

### Manual refinement of μ and classification of high confidence μ-SNPs and associated target genes

For each of the 36 unique candidate μ-SNP’s prioritized for follow-up, we visually examined the GRO-seq tracks within the surrounding genomic region in order to manually refine the bidirectional calls originally made by Tfit. We passed these manually refined regions of bidirectional transcription (Supp File 2) to a custom script that calculates the midpoint between the peak signal intensity in each half of the region, resulting in a new estimate of μ for each bidirectional relying both on manual refinement and computational analysis. We then averaged the μ values for any sets of replicates in which the specified bidirectional had robust signal and compared the refined μ estimate to the SNP location. SNPs located within 500 bp (+/−) of the refined μ locations were classified as high confidence μ-SNPs and examined further (Supp Table 3). A custom python script, in conjunction with bedtools (v. 2.28.0), was used to identify genes within 1 Mb (+/−) of each SNP and append results from differential gene expression analysis (Supp File 3) for easy comparison with the relevant μ-SNP bidirectional signature. To account for the short 30-minute GRO-seq treatment timepoint potentially missing true signal changes in longer gene transcripts, a second, “truncated” differential gene expression analysis was performed. Assuming a conservative polymerase processivity rate of 3 kb/min^70^, gene transfer format (GTF) annotation files were generated corresponding to the first 90 kb downstream of all UCSC RefGene-annotated gene TSSs (downloaded August 2019); genes smaller than 90 kb were retained in full. Reads were then counted across these GTF files using the featureCounts algorithm within the R (v. 3.6.1) package Rsubread (v. 3.10) and analyzed with DESeq2 (v. 1.26.0) for differential expression (Supp File 4). Genes for which significant expression changes were primarily evident in the truncated DESeq2 analysis, including *NR3C1*, which spans ∼150 kb, are asterisked in Supp Table 3; data presented in the Results section for *NR3C1* are from this truncated analysis.

### Cell culture and reagents

BEAS-2B transformed normal human airway epithelial cells (ATCC) were grown in DMEM with L-glutamine and 4.5 g/L glucose (Corning) containing 10% FBS (VWR) and 1% penicillin/streptomycin (pen/strep; Corning). Primary human airway smooth muscle (HASM) cells (provided by Dr. Reynold Panettieri, Rutgers Biomedical Health Sciences) were cultured in Ham’s F12 Medium with L-glutamine (Corning) containing 10% FBS and 1% pen/strep. All cells were maintained in 5% CO_2_ at 37°C.

Dexamethasone (dex; Sigma-Aldrich) was dissolved in sterile 100% ethanol (vehicle) and used at a final concentration of 100 nM. Recombinant Human TNF-alpha Protein (TNF) purchased from R&D Systems was diluted in sterile 1X Dulbecco’s phosphate buffered saline (DPBS) containing 0.1% bovine serum albumin (BSA) and used at a final concentration of 20 ng/ml. 2,3,7,8-tetrachlorodibenzo-p-dioxin (TCDD) obtained from Cambridge Isotope Laboratories was diluted in sterile DMSO (vehicle) and used at a final concentration of 10 nM. Wood smoke particles were obtained and prepared as described previously^68^ and used at a final concentration of 1 mg/ml.

### Micro-C

Three-dimensional chromatin conformation was assayed using the Micro-C protocol described by Hsieh et al^71^, with the following specifications. BEAS-2B cells were grown to confluence on 15 cm tissue culture plates and treated with a 1:1000 dilution of ethanol (vehicle) in fresh complete medium for 1 hr. Dual-crosslinking was sequentially performed *in situ* using freshly prepared 1% methanol-free formaldehyde for 10 min and disuccinimidyl glutarate (Thermo Scientific; reconstituted in DMSO) at a final concentration of 3 mM for 45 min at room temperature. Cells were scraped in PBS into conical tubes and pelleted by centrifugation, then snap frozen in a dry ice/ethanol bath for storage at −80°C. Each pellet was resuspended in Micro-C Buffer #1 (MB #1: 50 mM NaCl, 10 mM Tris-HCl-pH 7.5, 5 mM MgCl2, 1 mM CaCl2, 0.2% NP-40, 1X Protease Inhibitor Cocktail (Thermo Scientific)) and immediately divided equally into 2 replicate samples containing ∼10 million cells each prior to incubation on ice for 20 min to release intact nuclei. Chromatin was fragmented using Micrococcal Nuclease (New England Biolabs) at a final concentration of 2000 gel units/ml (1:1000 dilution) in MB #1 and incubating at 37°C for 20 min with shaking at 850 rpm. These conditions were pre-optimized in BEAS-2B cells to yield a range of chromatin fragmentation containing ∼90% mono-nucleosomes and ∼10% di-nucleosomes. Fragmented chromatin ends were repaired, blunted and biotinylated as described^71^using biotin-ATP and -CTP (obtained from Axxora) and crosslinked nucleosomes were ligated by T4 DNA Ligase (New England Biolabs) for 2.5 hr at room temperature. Fragment ends were cleaned with Exonuclease III (New England Biolabs) and crosslinks were reversed by overnight incubation in 2 mg/ml Proteinase K (Sigma) in 1% SDS at 65°C. Chromatin was then phenol:chloroform:isoamyl alcohol extracted, ethanol precipitated, treated with RNase A (Invitrogen), and purified with a DNA Clean and Concentrator-5 Kit (Zymo) prior to separation on a 3.5% NuSieve Agarose Gel (Lonza). The band containing the ligated di-nucleosomal DNA (∼250-400 bp) was excised from the gel and DNA purified for library preparation using a ZymoGel Purification Kit (Zymo). Biotinylated di-nucleosomal DNA was captured using DYNAL MyOne Dynabeads Streptavidin C1 (Invitrogen) and used as input for the NEBNext Ultra II DNA Library Prep Kit for Illumina (New England Biolabs), which includes standard procedures for end-repair, A-tailing, and adaptor ligation. Five cycles of initial library amplification employing KAPA HiFi HotStart ReadyMix (KAPA Biosystems) and Multiplex Oligos for Illumina, Index Primers Set 1 (New England Biolabs; for i7 barcoding) were followed by quantitative PCR assessment of 10% of the pre-amplified product to determine the lowest number of additional PCR cycles for sufficient final library amplification with minimal PCR duplication. Pre-amplified libraries underwent 8 additional cycles of amplification and were then size-selected and purified using Ampure XP Beads (Agencourt) at 0.9X volumes. Libraries were pooled and sequenced on an Illumina NovaSeq 6000 instrument using paired-end 2×150 bp reads by the Genomics Shared Resource at the University of Colorado Anschutz Medical Campus.

### Micro-C computational analysis

Paired-end Micro-C data were analyzed using the HiC-Pro^72^(v. 2.11.4) pipeline, which independently aligns R1 vs R2 reads to the hg38 reference genome using bowtie2 (v. 2.2.9) in end-to-end mode. Prior to inputting raw fastq data into HiC-Pro, we merged R1 and R2 reads from each replicate, respectively. HiC-Pro then filters for valid interactions within specified bins of the genome using samtools (v. 1.1), python (v. 2.7), and the python packages bx-python (v. 0.8.9), scipy (v. 1.2.3), pysam (v. 0.16.0.1) and pandas (v. 0.24.2) to generate raw contact counts between corresponding pairs of loci, output as a single coverage file. We configured HiC-Pro to remove all singleton, multimapped, and duplicate reads, and binned reads at 1 kb intervals, in accordance with the highest suggested resolution for Micro-C. To account for systematic biases inherent in chromatin conformation assays, contact counts were normalized using the iced normalization package (v. 0.5.7). Contacts were then filtered to remove those with a normalized count < 1.0 and were visualized as contact maps using the plotgardener library (v. 1.4.2) in R (v. 4.2.3).

### Cloning, transfection and luciferase assays

Enhancer reporter constructs were amplified from BEAS-2B genomic DNA by PCR, cloned into pCR2.1-TOPO (Life Technologies), and subsequently ligated into the pGL3-Promoter vector backbone (Life Technologies) using SacI/XhoI (p*COL8A1*) or KpnI/XhoI (all others). p*FKBP5*, p*TNFAIP3*, and p*MT2A* positive control constructs have been described^41,46,73^. The QuikChange II Site-directed Mutagenesis (SDM) Kit from Agilent Technologies was used as instructed by the manufacturer to generate reporter constructs harboring variant alleles (p*GCNT1* C>T, p*NR3C1* C>T, p*rs258753* T>C, p*rs864354* G>T). PCR primer sequences used for cloning and SDM and detailed information pertaining to each reporter construct and related μ-SNP are presented in Supp Table 4.

For transfection experiments, BEAS-2B cells were seeded on 96-well tissue culture plates at 20K cells/well in complete medium. The next day, cells were transfected with a total of 200 ng DNA/well (10:1 ratio of firefly luciferase reporter construct to *Renilla* luciferase internal control (pSV40-RL; Promega)) using Lipofectamine 2000 (0.5 ul/well) from Life Technologies. Transfection complexes were removed the following day and cells were treated in fresh complete medium for 8 hr prior to luciferase assay. HASM1 cells were seeded on 96-well tissue culture plates at 50K cells/well in complete medium. Medium was replaced two days later with fresh complete and the following day, cells were transfected with a total of 125 ng DNA/well (10:1:1 ratio of firefly luciferase to renilla luciferase to mCherry plasmid, respectively) using Lipofectamine 3000 (0.3 ul/well) and P3000 Reagent (0.8 ul/well) from Life Technologies. Transfection complexes were removed the following day and cells were treated in fresh complete medium for 24 hr prior to luciferase assay. HASM2 cells were seeded on 96-well plates at 40K cells/well in antibiotic-free medium. The next day, cells were transfected with 200 ng total DNA/well (10:1:1 ratio as described for HASM1 cells) using FuGENE6 (1 ul/well) from Promega. Transfection complexes were replaced the following day with fresh complete medium and cells allowed to recover an additional 24 hr prior to 24 hr treatment and luciferase assay.

Luciferase assays were performed using the Dual-Luciferase Reporter Assay System (Promega) and Infinite M1000 Plate Reader (Tecan) as previously described^74^, with the following modifications for cells grown in 96-well plates: each well was lysed in 25 ul 1X lysis reagent and luminescence detected from 5 ul lysate combined with 40 ul of each assay reagent. Each experiment was performed in biologic sextuplet (except for those employing TCDD, which were run in biologic quadruplicate to reduce waste generation) and repeated at least once with qualitatively similar results.

### RNA purification and quantitative RT-PCR (qRT-PCR)

BEAS-2B cells were grown to confluence in 6-well tissue culture plates and treated with vehicle (DMSO) or TCDD (10 nM) for 4 hr. Wells were rinsed 3X in DPBS and then fresh medium containing 100 nM dex was added to all wells for an additional 4 hr. Cells were harvested in TRIzol and RNA purified using the PureLink RNA Mini Kit from Life Technologies prior to qRT-PCR, performed with normalization to *RPL19* as previously described^74^. Primers used for qRT-PCR are described in Supp Table 5.

### Statistical notes

Statistical tests for comparing reporter assays and RT-qPCR data are described in the Figure Legends. In all cases, tests were performed under the assumption of normality of the underlying distributions.

### Data availability statement

Micro-C data has been submitted to GEO and the accession number is pending. All genomics data used in this manuscript are publicly available, with accession numbers identified above in the relevant methods sections. Information for access to GERA data can be found at: https://divisionofresearch.kaiserpermanente.org/research/research-program-on-genes-environment-and-health/

## Data Availability

All data produced in the present work are contained in the manuscript or are available upon reasonable request.

